# Topical 30% Ascorbic Acid in Dimethyl Sulfoxide for the Treatment of Cutaneous Squamous Cell Carcinoma *In Situ*

**DOI:** 10.1101/2025.10.13.25337920

**Authors:** Patricia Miller, Jon Ward, Michael Morgan, Briant Burke

## Abstract

**Background:** Cutaneous squamous cell carcinoma in situ (SCCIS) is a common nonmelanoma skin cancer with rising incidence and substantial treatment costs. While Mohs micrographic surgery remains the standard of care, cost, accessibility, and cosmetic concerns limit its use for some patients. Topical alternatives such as imiquimod and 5-fluorouracil are associated with variable efficacy and frequent adverse effects.

**Objective:** To evaluate the efficacy and tolerability of a 30% ascorbic acid (vitamin C) solution in 95% dimethyl sulfoxide (DMSO) for the treatment of biopsy-confirmed SCCIS.

**Methods:** In this open-label study, 17 patients with 27 histologically confirmed SCCIS lesions applied 30% ascorbic acid in DMSO topically twice daily for 12 weeks. Lesion size and histologic resolution were assessed by repeat biopsy at week 12. Primary outcome was complete resolution of SCCIS; secondary outcomes included lesion size reduction and adverse events.

**Results:** Complete histologic resolution occurred in 15 of 27 lesions (56%), while 44% of lesions showed >85% reduction in size. The mean reduction in lesion area was 71%. Only 2 lesions (7%) failed to respond. Most residual lesions demonstrated actinic atypia without carcinoma and were successfully treated with cryotherapy. No patients discontinued due to adverse effects. Ninety-four percent of participants (16/17) avoided surgical excision.

**Conclusions:** Topical 30% ascorbic acid in DMSO demonstrated promising efficacy and excellent tolerability in the treatment of SCCIS, offering a potential noninvasive alternative to surgery. Larger, controlled trials are warranted.

## INTRODUCTION

Skin cancer is the most common malignancy in the United States and among fair-skinned populations worldwide, with more cases diagnosed each year than all other cancers combined.^1^,^2^ Basal cell carcinoma (BCC) is the most frequent subtype, followed by cutaneous squamous cell carcinoma (SCC), which accounts for approximately 1.8 million cases annually in the United States.^3^ The annual cost of treating SCC is estimated at $4.2 billion, representing a substantial public health and economic burden.^4^ Current consensus guidelines recommend Mohs micrographic surgery as the treatment of choice for many SCC lesions.^5^ However, cost, limited accessibility, and concerns regarding cosmetic outcomes and postoperative scarring, particularly on the face, remain significant drawbacks.^5^

Topical therapies, including the antineoplastic agent 5-fluorouracil (5-FU) and the immune-response modifier imiquimod, have been employed as non-surgical alternatives. Although 5-FU is FDA-approved for actinic keratosis and superficial BCC, it is used off-label for SCC in situ (SCCIS). Imiquimod is FDA-approved for SCCIS. Reported efficacy rates for these topical treatments range from 35–75%, and both are associated with considerable local irritation and inflammation.^6^ Imiquimod may produce persistent hypopigmentation in up to 80% of treated sites,^7^ while 5-FU commonly induces burning, pruritus, and pain, often resulting in early discontinuation.^8^ Consequently, there is a need for an effective, well-tolerated, and cosmetically favorable topical therapy for SCCIS.

Ascorbic acid (vitamin C) has a long, controversial history as an antineoplastic agent.^9^ Renewed interest has emerged following evidence that high-dose intravenous ascorbate can generate cytotoxic hydrogen peroxide within tumor microenvironments, selectively affecting malignant cells while sparing normal tissue.^10-12^ Building on this mechanistic rationale, Burke and Bailie previously demonstrated the clinical efficacy of a 30% ascorbic acid solution in 95% dimethyl sulfoxide (DMSO) for the topical treatment of BCC, achieving lesion resolution rates exceeding 80% within 8 weeks.^13^

Given the biochemical and histologic similarities between BCC and SCCIS, we hypothesized that topical ascorbic acid might also induce regression of SCCIS through redox-mediated cytotoxicity. DMSO, a well-characterized skin-penetration enhancer, facilitates transdermal delivery of hydrophilic compounds, potentially augmenting intralesional vitamin C concentration and therapeutic effect.

The present study evaluates the efficacy, safety, and tolerability of 30% ascorbic acid in 95% DMSO for the topical treatment of biopsy-proven SCCIS.

### Study Rationale

SCCIS represents a therapeutic niche in which current topical and surgical options are either cosmetically burdensome or poorly tolerated. Given prior evidence supporting high-dose ascorbic acid as a redox-active antineoplastic agent and its demonstrated efficacy in topical BCC therapy, this study investigates whether a 30% ascorbic acid formulation in dimethyl sulfoxide (DMSO) could provide a similarly effective, well-tolerated, and noninvasive treatment for SCCIS. The goal was to assess clinical and histologic outcomes while further elucidating the potential of ascorbate-based topical therapy as a cost-effective adjunct or alternative to surgery.

## METHODS

### Study Design

This was an open-label, prospective clinical trial evaluating the efficacy and tolerability of a 30% (w/v) ascorbic acid (vitamin C) solution in 95% (v/v) dimethyl sulfoxide (DMSO) for the topical treatment of biopsy-confirmed cutaneous squamous cell carcinoma in situ (SCCIS). The study was conducted in accordance with institutional guidelines for clinical research and the principles of the Declaration of Helsinki (2013 revision). The protocol was registered on ClinicalTrials.gov (Identifier NCT04279535).

### Participants

Patients were recruited from community dermatology practices. Eligible participants were adults (≥18 years) with histologically confirmed, previously untreated SCCIS. Exclusion criteria included current treatment for any malignancy, a history of non-cutaneous cancer, or inability to comply with study procedures. Detailed written informed consent was obtained from each participant prior to enrollment.

Sixty-three patients were screened for eligibility (Figure 1). Forty-four were excluded— 11 for not meeting inclusion criteria and 33 for declining participation—resulting in 19 patients allocated to treatment. Two were lost to follow-up for unknown reasons, yielding 17 patients who completed the 12-week protocol and were included in the analysis.

**Figure 1.**
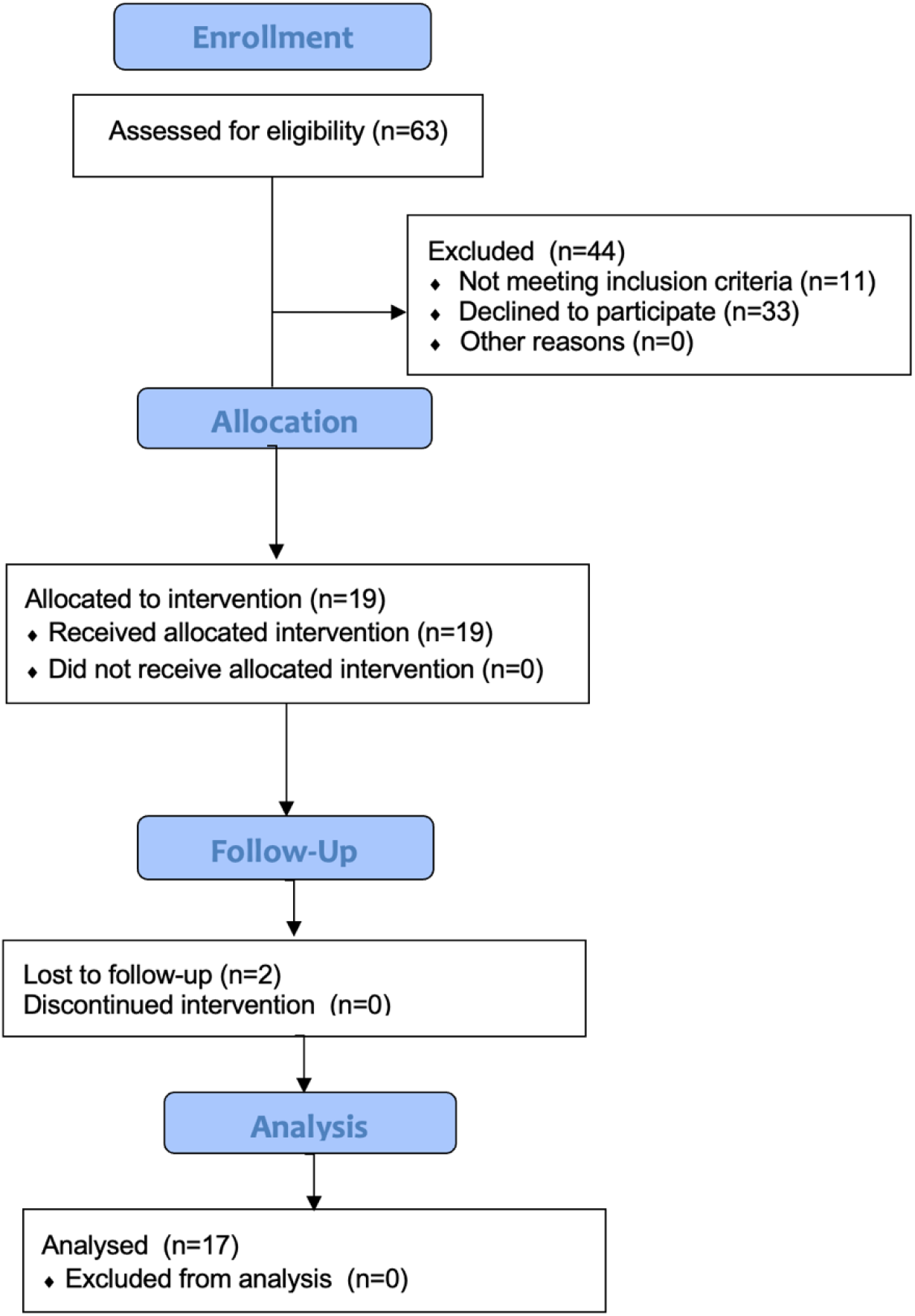
CONSORT flow diagram summarizing patient screening, enrollment, allocation, and analysis.

### Intervention

Participants applied a topical 30% (w/v) ascorbic acid solution prepared in 95% (v/v) DMSO and 5% (v/v) distilled water. A small cuticle brush was provided for home application. Each brush stroke delivered approximately 23 µL of solution—sufficient to moisten the lesion surface while minimizing contact with adjacent skin. Patients were instructed to apply the formulation twice daily for 12 weeks or until clinical clearance was observed.

Follow-up visits occurred at baseline, 4 weeks, 8 weeks, and 12 weeks (±2 weeks). At the final visit, each lesion underwent repeat shave biopsy to assess histologic response. Patient adherence and adverse events were assessed at each visit.

### Outcome Measures

The **primary endpoint** was complete histologic resolution of SCCIS at week 12, defined as absence of carcinoma on repeat biopsy.

**Secondary endpoints** included:

1. Percent reduction in lesion area (calculated from measured longest perpendicular diameters),
2. Presence of residual atypia or actinic keratosis,
3. Incidence and severity of local or systemic adverse effects, and
4. Need for subsequent procedural treatment.

### Statistical Analysis

Descriptive statistics were used to summarize demographic and clinical data. Continuous variables are expressed as mean ± standard error of the mean (SEM), and categorical variables as counts and percentages. Changes in mean lesion size from baseline to week 12 were evaluated using paired comparisons of group means. Given the small sample size, results were interpreted descriptively rather than inferentially.

### Ethical Approval

The study was approved by the appropriate institutional review committee and complied with all applicable regulatory and ethical requirements. All participants provided written informed consent prior to participation.

## RESULTS

### Patient Enrollment and Characteristics

A total of 63 patients were assessed for eligibility (**Figure 1**). Of these, 44 were excluded—11 who did not meet inclusion criteria and 33 who declined to participate. Nineteen participants were enrolled, of whom two were lost to follow-up for unknown reasons, leaving 17 patients who completed the study and were included in the analysis. No patient discontinued treatment due to adverse effects.

Participants ranged in age from 41 to 78 years (mean 64.5 years); seven were women and ten were men. All patients had biopsy-confirmed SCCIS lesions prior to enrollment. Lesion sites included the scalp (n = 5), face (n = 3), forehead (n = 3), nose (n = 3), neck (n = 3), ear (n = 2), hand (n = 4), and arm (n = 4).

### Lesion Response and Histologic Outcomes

Twenty-seven SCCIS lesions were treated across 17 patients. After 12 weeks of twice-daily topical application of 30% ascorbic acid in DMSO, histologic clearance was achieved in 15 of 27 lesions (56%). The remaining lesions demonstrated variable residual atypia or transformation to actinic keratosis morphology. Mean lesion size decreased from 1.06 ± 0.18 cm^2^ at baseline to 0.30 ± 0.09 cm^2^ at 12 weeks, representing an average 71% reduction (p < 0.05, descriptive). Twelve lesions (44%) exhibited > 85% size reduction, and only 2 lesions (7%) failed to respond.

Most residual atypical lesions were subsequently treated with cryotherapy. Overall, 16 of 17 patients (94%) avoided surgical excision. Microscopic examination of post-treatment specimens revealed focal residual atypia and increased lymphocytic infiltration consistent with treatment response (**Figure 2**). Perilesional inflammation was minimal, except in one patient whose final biopsy was delayed by two weeks to allow resolution of transient erythema.

**Figure 2.**
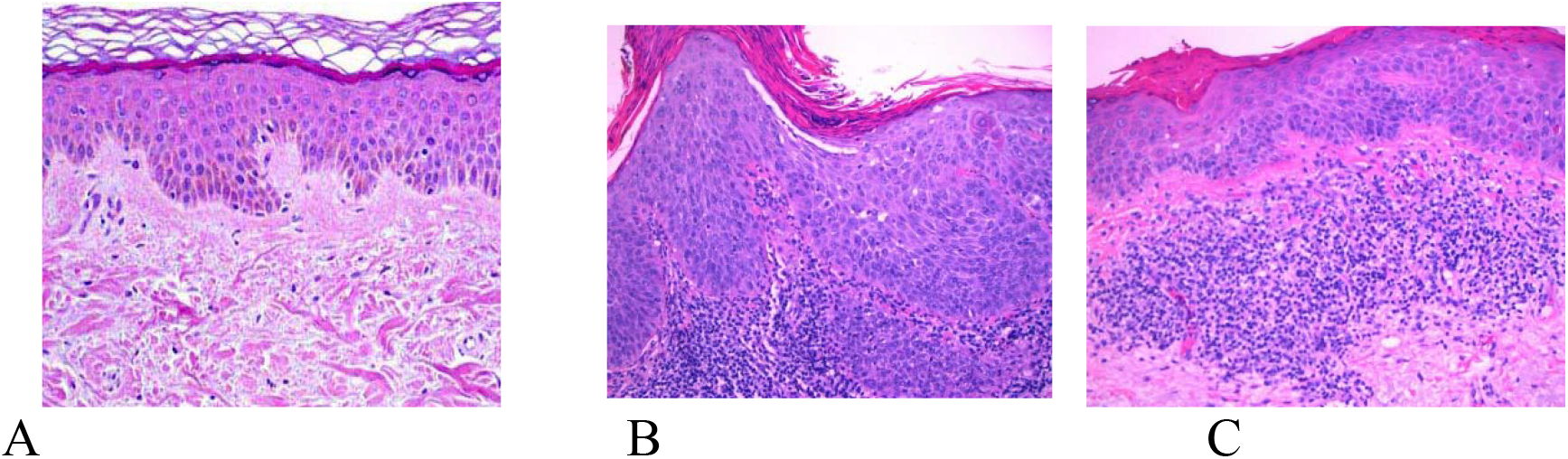
Representative histopathology of SCCIS before and after treatment with 30% ascorbic acid in DMSO for 12 weeks. (A) Normal skin; (B) Pretreatment SCCIS showing full-thickness atypia; (C) Post-treatment specimen demonstrating resolution of carcinoma with residual atypia and lymphocytic infiltrate.

#### Safety and Tolerability

No participants reported pain, pruritus, burning, or ulceration during therapy. Local reactions were limited to mild erythema at treated sites. There were no systemic adverse effects and no cases of hypopigmentation, a common sequela observed with imiquimod.

Lesion size reduction followed a time-dependent response consistent with an exponential or “log-kill” kinetic pattern characteristic of many antineoplastic therapies (**Figure 3**).

**Figure 3.**
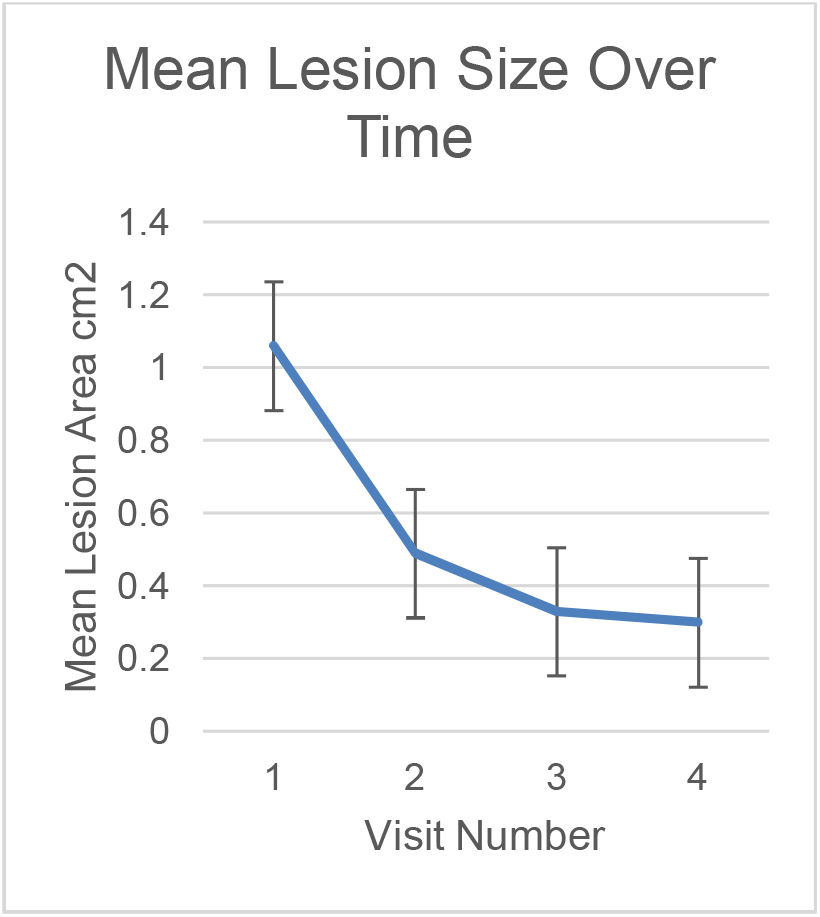
Mean percentage reduction in lesion area over 12 weeks of treatment with 30% ascorbic acid in DMSO.

## DISCUSSION

### Summary of Findings

Topical 30% ascorbic acid in 95% DMSO produced meaningful clinical and histologic improvement in cutaneous squamous cell carcinoma in situ (SCCIS). More than half of treated lesions achieved complete histologic clearance, with a mean lesion-size reduction of 71%. Nearly all patients (94%) avoided surgical excision, and the formulation was well tolerated without reports of irritation or pain. These findings suggest that concentrated topical ascorbic acid may represent a practical, noninvasive therapeutic option for selected SCCIS lesions.

### Comparison with Existing Therapies

Current nonsurgical treatments for SCCIS—such as 5-fluorouracil (5-FU) and imiquimod—are limited by variable efficacy and frequent local adverse effects.^6-8^ In contrast, the present formulation was well tolerated, with no pain, pruritus, or hypopigmentation, a complication reported in up to 80% of imiquimod-treated sites.^7^ This improved tolerability may enhance adherence and patient satisfaction, especially for cosmetically sensitive areas.

Although the overall clearance rate (56%) was lower than that reported for the same formulation in basal cell carcinoma (BCC) (>80% resolution within 8 weeks),^13^ SCCIS represents a more advanced keratinocytic dysplasia with greater metabolic heterogeneity.^20^ This may explain the persistence of residual atypia in many lesions. Nevertheless, the observed downgrading from carcinoma in situ to actinic keratosis in most cases is clinically meaningful and supports the use of high-dose ascorbate to reduce surgical morbidity.

### Proposed Mechanism of Action

High-dose ascorbate functions as a pro-oxidant within the tumor microenvironment, generating hydrogen peroxide (H_2_O_2_) through redox cycling in the presence of catalytic transition metals.^15-17^ Elevated labile ferric iron in malignant tissue promotes this reaction, producing selective oxidative stress that preferentially damages tumor cells.^16,17^ Normal keratinocytes possess higher catalase activity and other antioxidant defenses that neutralize H_2_O_2_^21,22^.

Histopathologic findings in this study—marked reduction of carcinoma with residual atypia—support a metabolic-threshold model in which transformed SCCIS cells, characterized by decreased catalase expression, are selectively vulnerable to peroxide-mediated apoptosis.^20,21^ Actinic keratosis cells may retain sufficient enzymatic defenses to survive. This mechanism aligns with evidence that pharmacologic ascorbate targets metabolically reprogrammed cells while sparing normal tissue.^16,17^.

### Role of Dimethyl Sulfoxide (DMSO)

DMSO is a well-characterized skin-penetration enhancer that transiently disrupts stratum-corneum lipid organization to facilitate transdermal drug delivery.^25^ It also exhibits mild anti-inflammatory and differentiating properties in certain neoplastic cell lines.^26,27^ However, topical DMSO alone has shown little direct antitumor activity.^28,29^ In the present study, DMSO most likely served as a carrier, improving dermal penetration of ascorbate and accelerating clinical response compared with aqueous formulations, which have required ≥22 weeks for visible regression.^23,24^

### Ascorbate Chemistry and Pharmacologic Kinetics

The chemistry of ascorbate in aqueous and aprotic environments further informs its biological activity. In water, equilibrium exists among ascorbic acid, the ascorbyl radical, and dehydroascorbate, all of which participate in redox cycling and hydrogen-peroxide generation.^31^ In contrast, DMSO, an aprotic solvent, alters hydrogen bonding and redox dynamics, potentially enhancing intracellular diffusion of reduced ascorbate species. The combination of DMSO with photodynamic therapy has been shown to improve dermal penetration of 5-aminolevulinic acid for nonmelanoma skin cancers, supporting its role as an effective carrier.^28,29^ The lesion-regression curve observed here followed an exponential or “log-kill” pattern typical of pharmacologic tumor-cell-death kinetics under sustained cytotoxic exposure.^30^ These findings suggest that both solvent effects and redox kinetics contribute to therapeutic efficacy.

### Clinical Implications

SCCIS rarely undergoes spontaneous regression, and surgery remains the standard of care. However, for patients with comorbidities or lesions in cosmetically critical areas, a safe topical alternative could reduce morbidity and cost. The present data indicate that high-concentration ascorbic acid can shrink lesions, downgrade histology, and potentially serve as a neoadjuvant before excision. Its absence of discomfort or pigment alteration underscores its cosmetic advantage.

### Study Limitations

Limitations include small sample size and an open-label design which therefore lacked a placebo control. Although DMSO alone was not evaluated, prior reports of aqueous ascorbate efficacy in BCC and SCC suggest that the antineoplastic activity stems from ascorbate itself.^23,24^ The cohort lacked ethnic diversity, and long-term recurrence data remain pending.

### Future Directions

Larger randomized trials are warranted to confirm efficacy, optimize concentration and duration, and further explore mechanistic biomarkers such as catalase expression and redox balance.

## Conclusions

Management of SCCIS depends on factors such as lesion size, anatomic site, and cosmetic considerations. A 30% ascorbic acid formulation in DMSO achieved significant lesion regression and histologic improvement while allowing nearly all patients to avoid surgery. The treatment was inexpensive, well tolerated, and cosmetically favorable, with no pain, irritation, or pigmentary change. Given its simplicity and low cost, topical ascorbic acid may represent a feasible first-line or adjunctive therapy for SCCIS, particularly for cosmetically sensitive regions or in patients who are poor surgical candidates. Larger, controlled studies are needed to validate these findings and establish optimal treatment parameters.

## Author Contributions (CRediT Statement)

**Briant Burke, MD, MS:** Conceptualization, Methodology, Formal Analysis, Writing – Original Draft, Supervision

**Patricia Miller, MD:** Data Collection, Validation, Writing – Review & Editing

**Jon Ward, MD:** Project Administration, Writing – Review & Editing

**Michael Morgan, MD:** Histologic Analysis, Visualization, Review & Editing

All authors have read and approved the final manuscript.

## Ethical Approval

The study was conducted in accordance with the ethical standards of the institutional research committee and with the 2013 revision of the Declaration of Helsinki. All participants provided written informed consent prior to enrollment.

## Data Availability Statement

De-identified data supporting the findings of this study are available from the corresponding author upon reasonable request.

## Acknowledgments

The authors thank the participating dermatology clinics and staff for their assistance with patient recruitment and follow-up.

